# Longitudinal Cerebrospinal Fluid Biomarker Changes Following Shunt Surgery in Idiopathic Normal Pressure Hydrocephalus - Insights from the Gothenburg POiNT study

**DOI:** 10.64898/2026.11.11.26366969

**Authors:** Daniel Jaraj, Kerstin Andrén, Per Hellström, Katarina Laurell, Henrik Zetterberg, Carsten Wikkelsö, Mats Tullberg

## Abstract

**Background:** The underlying pathophysiology of idiopathic normal pressure hydrocephalus (iNPH) is incompletely understood and reliable predictors of shunt response are lacking. Cerebrospinal fluid (CSF) biomarkers may provide insights into disease mechanisms and help identify patients most likely to benefit from surgery. The aim of this study was to explore CSF biomarker dynamics following shunt surgery and their relationship with clinical outcome.

**Methods:** The POiNT (Prediction of Outcome in iNPH) was a prospective longitudinal study. One hundred and thirty-five patients with iNPH underwent standardized clinical assessment and lumbar CSF sampling 3.6 (SD 2.1) months before and 5.7 months (SD 2.4) after shunt surgery. CSF was analyzed for biomarkers of astrogliosis, axonal injury, amyloid, and tau pathology, and synaptic dysfunction. Clinical outcomes were assessed using the iNPH scale. Associations between baseline biomarker levels, postoperative biomarker changes, and clinical outcomes were evaluated using adjusted correlation and regression analyses.

**Results:** In total, 69% (n=93) improved after shunt surgery. Substantial postoperative increases were observed for most CSF biomarkers: The largest relative increases were seen for growth-associated protein 43 (GAP43) (2.92-fold change), followed by Phosphorylated tau (2.65-fold change), Neurogranin and total-tau (both 2.29-fold change) as well as Aβ40 (1.93-fold change), and Aβ42 (1.76-fold change) (all p<0.001). In adjusted analyses, postoperative change in several biomarker levels were associated with degree of clinical improvement. However, baseline biomarker levels generally showed limited predictive value. Lower baseline monocyte chemoattractant protein-1 (MCP1) levels were associated with greater odds of overall clinical improvement but had a modest predictive performance (AUC 0.67). No baseline biomarker demonstrated strong discrimination of treatment response.

**Conclusion:** Shunt surgery was associated with substantial changes in CSF biomarker concentrations, several of which correlated with the degree of clinical improvement. Although baseline levels did not predict clinical outcome, the observed biomarker changes may reflect biological responses to treatment, altered CSF dynamics, or both .iNPH. Future studies examining these processes relating to disease reversibility might help elucidate pathophysiological mechanisms in iNPH.

## INTRODUCTION

Despite major scientific advances in the field of idiopathic normal pressure hydrocephalus (iNPH) in recent years, several important knowledge gaps remain. Currently, two of the most important challenges are a lack of understanding of the underlying pathophysiology and a need to find robust predictors of shunt response (1). Clarifying the relationship between cerebrospinal (CSF) biomarkers, and their association with treatment response may not only improve patient selection for surgery but also provide a window into the biological mechanisms that underlie symptom development and reversibility in iNPH (2).

So far, the predictive performance of CSF biomarkers, reported in the literature, has been inconsistent and most studies have relied on cross-sectional designs leaving the longitudinal dynamics of CSF biomarkers following shunt surgery incompletely characterized (3).

Consequently, the extent to which biomarker changes reflect treatment-related biological effects, disease reversibility, or mechanisms underlying clinical improvement remain unclear.

The aim of this study was therefore to examine CSF biomarkers in patients with iNPH undergoing shunt surgery, to assess longitudinal changes, their association with clinical improvement and capacity to predict outcome of shunt surgery.

## METHODS

### Design and study sample

The study sample has been described in detail previously (4, 5). This was a prospective study conducted at two Swedish centers: The Sahlgrenska University Hospital in Gothenburg and Östersund Hospital, with consecutive enrollment from 1 January 2014 to 31 December 2016. A total of 272 patients were investigated for suspected iNPH of which 177 were diagnosed with iNPH. Of these, 160 patients underwent shunt surgery and 135 had complete baseline and follow-up data including pre- and postoperative lumbar CSF samples and were included in the present study.

### Clinical examinations

All patients underwent a series of clinical and radiological examinations by a multidisciplinary team specializing in disorders of CSF dynamics. These examinations included clinical assessment by a neurologist, a neuropsychologist, and a physiotherapist, a standard MRI of the brain and lumbar puncture for assessment of opening pressure and collection of CSF. Clinical symptoms and outcomes were rated on the Gothenburg iNPH scale introduced by Hellström et al. (6), where the total score is the mean of four domain scores: gait, balance, cognitive function, and continence - each transformed to a 0-100 metric with higher scores indicating better function, and the gait domain given double weight. Outcome was defined as postoperative score minus preoperative score (delta iNPH scale score). A postoperative increase in the iNPH scale score of five points or more defined clinical improvement (6). Functional disability was rated with the modified Rankin Scale (mRS), (0-5: higher scores indicating lower level of function) (7) and overall cognitive function by the Mini-Mental State Examination (MMSE) (0-30: higher scores indicating better cognitive function)(8).

All examinations were made in accordance with a standardized protocol, and all clinical variables were assessed identically before- and after shunt surgery. The diagnosis of iNPH was made in accordance with international guideline criteria (9).

The decision for surgery was made at a multidisciplinary conference, where all examinations were reviewed together with a neurosurgeon. CSF-dynamic testing (with a CSF-tap test or lumbar infusion study) was done in selected cases when the diagnosis was considered uncertain, mostly due to indistinct or atypical symptomatology.

### CSF biomarker procedures

Lumbar puncture was performed a mean of 3.6 months (SD 2.1) before shunt surgery and a mean of 5.7 months (SD 2.4) postoperatively using standard procedures. The opening pressure was recorded, and CSF samples were collected, aliquoted and frozen at -80°C until analysis. All analyses were done at the Neurochemistry Laboratory at Sahlgrenska University Hospital, by board-certified laboratory technicians. Amyloid β (Aβ) - isoforms (Aβ40, Aβ42) and soluble amyloid precursor protein α (sAPPα) and β (sAPPβ) and monocyte chemoattractant protein 1 (MCP1) were analyzed by electrochemiluminescence assays (Meso Scale Discovery, Rockville, MD, USA). Validated in- house ELISA methodology was used to analyze neurofilament light protein (NfL) (10), neurogranin (NG) (11), GAP43 (12) and GFAP (13), whereas CSF levels of T-tau, and P-tau were measured using commercially available Lumipulse technology (Fujirebio, Ghent, Belgium), as previously described (14). Chitinase-3-like protein 1 (YKL40) was measured using Human Chitinase3-like 1 Quantikine ELISA Kit (R&D Systems, Minneapolis, MN) (14). All concentrations are given in ng/L. All samples were analyzed in one round of experiments using one batch of reagents. Intra- assay coefficients of variation were monitored using internal quality control samples.

### Treatment

The patients were treated with CSF diversion using a ventriculoperitoneal (n=131) or ventriculoatrial (n=4) shunt: PS Medical Strata, Medtronic (Goleta, California, USA). Valves with an antisiphon device (ASD) and a setting of 1.5 were used in all cases.

Postoperatively, shunt function was assessed by evaluation of clinical symptoms and CT or MRI. If doubts regarding shunt patency remained following computed tomography (CT) or magnetic resonance imaging (MRI), a radionuclide shuntography (15) or a lumbar infusion test was made.

After evaluation of shunt patency, patients underwent standardized reassessment of symptoms and clinical findings. Patients were deemed improved if their postoperative iNPH scale score had increased by at least five points (6).

### Patient consent and ethical considerations

The study was approved by the Regional Ethical Review Board in Gothenburg (Dnr 328-14, T439- 15) in accordance with the Declaration of Helsinki. Written informed consent was given by the patients or their next-of-kin.

### Statistical analyses

Descriptive statistics are reported as means and standard deviations (SDs) for clinical outcomes and as geometric means with coefficients of variation (CVs) for biomarker values. Statistical analyses were performed using parametric methods. All biomarkers were log-transformed prior to analysis to address skewness.

Changes in the clinical outcomes from pre- to post-surgery were analyzed using a paired t test. Changes in cerebrospinal fluid (CSF) biomarker levels were analyzed using a fixed-effects regression model for repeated measures, with an unstructured covariance matrix. Fold changes (geometric mean ratios) were derived by exponentiating the estimated mean difference on the log scale.

Associations between baseline biomarker levels and changes in clinical outcomes were evaluated using Pearson correlation coefficients. Adjusted analyses were conducted using partial Pearson correlations, controlling for age and iNPH scale total score at baseline. Associations between changes in biomarker levels and clinical improvement were assessed similarly.

To examine the predictive value of baseline biomarker levels for clinical improvement, logistic regression analyses were performed. Adjusted models (controlling for age and iNPH scale total score at baseline) were fitted using log-transformed biomarker values, with results presented as odds ratios (ORs) of improvement versus no improvement per 50% increase in biomarker level. Predictive performance was evaluated by calculating the area under the receiver operating characteristic (ROC) curve (AUC). Missing data were handled using multiple imputation by chained equations under the assumption of missing at random.

All statistical tests were two-sided and conducted at the 5% significance level. Statistical analyses were conducted using SAS/STAT® software, version 9.4 (SAS Institute Inc, Cary, NC, USA. Corp.).

## RESULTS

### Clinical outcomes

Baseline demographics are shown in table 1. Among the 135 included patients 69% (n=93) improved after shunt surgery, and significant clinical improvement was observed across all domains of the iNPH scale (Table 2). The mean iNPH total score increased from 53.8 to 66.2 points (p<0.001).

**Table 1.** Demographic data at baseline

|  | All (n=135) | Improved (n=93) | Unimproved (n= 42) |
|---|---|---|---|
| Age, years; mean (SD) | 74 (7) | 73 (7) | 75 (7) |
| Sex (men) n, % | n=89, 66% | n=60, 65% | n=29, 69% |
| BMI, kg/m<sup>2</sup>;mean (SD) | 27 (4) | 27 (4) | 27 (5) |
| Duration of symptoms, months; mean (SD) | 40.0 (32.0) | 39.2(31.5) | 41.7 (33.0) |
| Time from baseline LP to surgery, months; mean (SD) | 3.6 (2.1) | 3.6 (2.0) | 3.7 (2.4) |
| Time from surgery to follow-up, months; mean (SD) | 5.7 (2.4) | 5.5 (2.3) | 6.0 (2.7) |
| Comorbidity | n=92, 68% | n=63, 67% | n=29, 69% |
| Heart disease | n=41, % | n=27 | n=14 |
| Hypertension | n=78 | n=54 | n=24 |
| Diabetes Mellitus | n=40 | n=29 | n=11 |
| MMSE | 26 (24-28) | 26 (24-28) | 26 (23-28) |
| iNPH-scale preoperative; mean (SD) | 53.8 (17.8) | 53.0 (17.8) | 56.0 (18.0) |
| iNPH-scale postoperative; mean (SD) | 66.2 (18.2) | 70.1 (17.2) | 57.5 (17.4) |

**Abbreviations:** SD: Standard Deviation, MMSE: Mini Mental State Examination, BMI: Body Mass Index, LP: Lumbar puncture

**Table 2.**
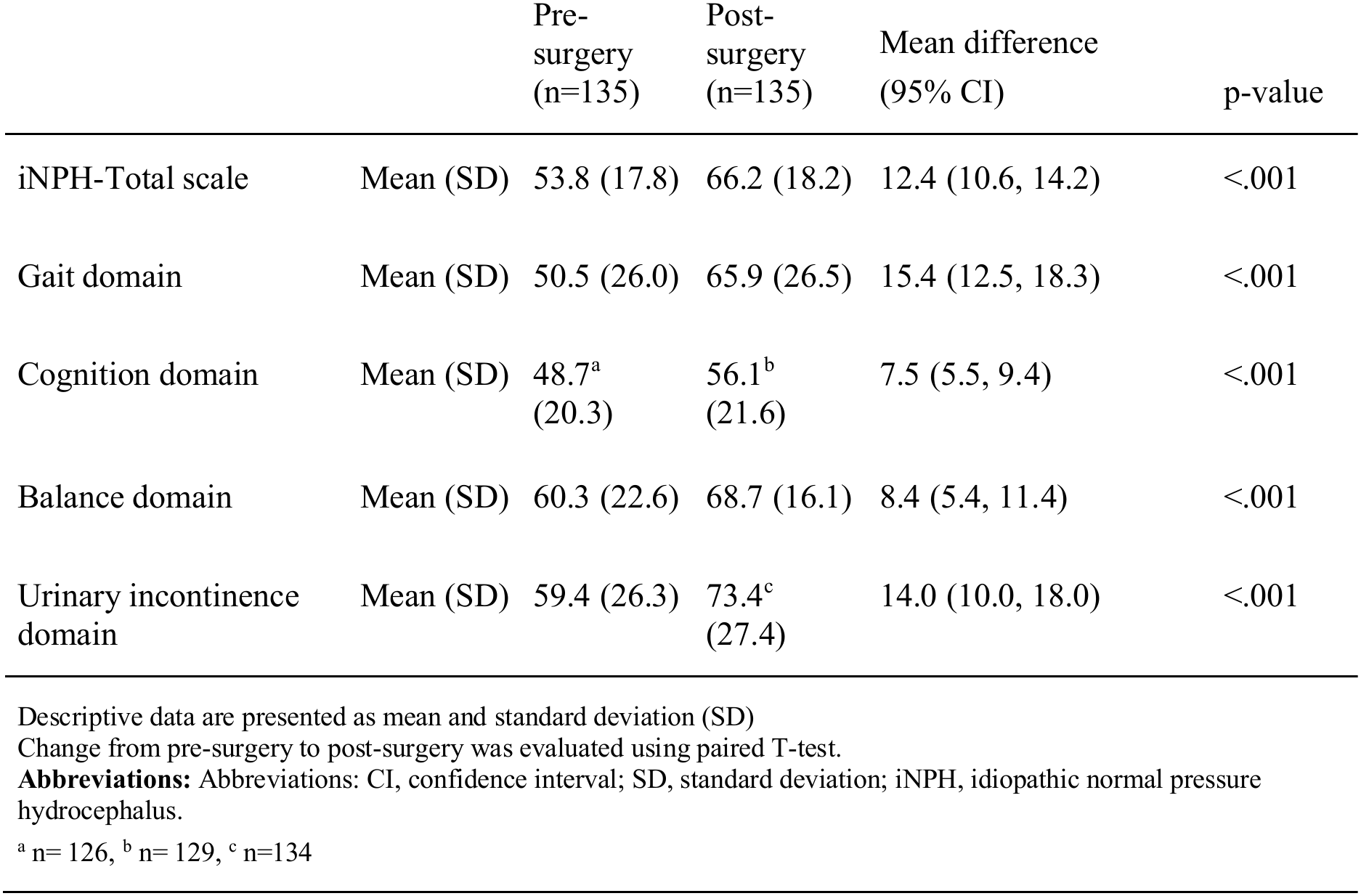
Change in iNPH total scale from pre- to post-shunt surgery

### Changes in CSF biomarker concentrations following shunt surgery

Substantial postoperative changes in CSF biomarker concentrations were observed (Table 3). All tested biomarkers increased significantly after surgery, with the largest relative increase observed for GAP43 (2.92-fold change), followed by P-tau (2.65-fold change), NG and T-tau (both 2.29-fold change) as well as Aβ40 (1.93-fold change), and Aβ42 (1.76-fold change) (all p<0.001). Because Aβ40 increased more, relative to Aβ42, the ratio between these decreased slightly.

**Table 3.** Changes in CSF biomarkers from pre- to post-surgery.

|  |  | Pre-surgery<br>(n=135) | Post-surgery<br>(n=98) | Fold change (95% CI) | p-value |
|---|---|---|---|---|---|
| Aβ40 | GeoMean (CV) | 6120 (0.38) | 11890 (0.40) | 1.93 (1.79, 2.07) | <.001 |
| Aβ42 | GeoMean (CV) | 483 (0.41) | 876 (0.58) | 1.76 (1.61, 1.93) | <.001 |
| Aβ 42/40 ratio | GeoMean (CV) | 0.79 (0.27) | 0.74 (0.40) | 0.91 (0.87, 0.94) | <.001 |
| P-tau | GeoMean (CV) | 25.92 (0.50) | 67.54 (0.65) | 2.65 (2.40, 2.92) | <.001 |
| GFAP | GeoMean (CV) | 736 (0.48) | 855 (0.49) | 1.17 (1.09, 1.26) | <.001 |
| NFL | GeoMean (CV) | 1780 (0.65) | 2760 (0.57) | 1.57 (1.42, 1.73) | <.001 |
| T-Tau | GeoMean (CV) | 240 (0.60) | 535 (0.60) | 2.29 (2.05, 2.55) | <.001 |
| APPa | GeoMean (CV) | 137 (0.53) | 228 (0.55) | 1.66 (1.54, 1.80) | <.001 |
| APPß | GeoMean (CV) | 305 (0.47) | 474 (0.51) | 1.56 (1.45, 1.68) | <.001 |
| YKL40 | GeoMean (CV) | 148 (0.34) | 239 (0.43) | 1.60 (1.51, 1.70) | <.001 |
| MCP1 | GeoMean (CV) | 420 (0.27) | 436 (0.27) | 1.05 (1.01, 1.09) | 0.021 |
| NG | GeoMean (CV) | 94.53 (0.41) | 215 (0.47) | 2.29 (2.09, 2.51) | <.001 |
| GAP43 | GeoMean (CV) | 2160 (0.39) | 6240 (0.64) | 2.92 (2.59, 3.29) | <.001 |

Descriptive data are presented as geometric means (GeoMean) with coefficients of variation (CV).
Changes from pre-surgery to post-surgery were evaluated using a repeated measures regression model with an unstructured covariance matrix, accounting for missing data.
Biomarkers were log-transformed prior to analysis to account for skewness.
Results are presented as fold changes, calculated by exponentiating the mean change on the log scale.
**Abbreviations:** Aβ40, amyloid-ß 1–40; Aβ42, amyloid-ß 1–42; APPa, amyloid precursor protein alpha; APPß, amyloid precursor protein beta; CI, confidence interval; CV, coefficient of variation; P-tau, phosphorylated tau; GAP43, growth-associated protein 43; GFAP, glial fibrillary acidic protein; MCP1, monocyte chemoattractant protein-1; NFL, neurofilament light chain; NG,Neurogranin; T-Tau, total tau protein; YKL40, chitinase-3-like protein 1.

### Associations between biomarker changes and clinical improvement

Changes in several biomarkers were associated with postoperative clinical improvement (Table 4). In adjusted analyses, increases in GFAP, YKL40, and GAP43 were associated with greater improvement in the total iNPH scale score (partial r=0.20–0.28; all p<0.05).

**Table 4.** Correlation between change in biomarker levels and change in iNPH total scale score from pre- to post-surgery.

| Biomarker | Pearson correlation (95% CI) | p-value | Partial Pearson correlation* (95% CI) | p-value |
|---|---|---|---|---|
| Aβ40 | 0.25 (0.05, 0.43) | 0.017 | 0.21 (-0.005, 0.40) | 0.056 |
| Aβ42 | 0.25 (0.06, 0.42) | 0.011 | 0.17 (-0.03, 0.35) | 0.096 |
| Aβ 42/40 ratio | 0.05 (-0.17, 0.25) | 0.67 | -0.02 (-0.24, 0.20) | 0.85 |
| P-tau | 0.18 (-0.02, 0.37) | 0.071 | 0.18 (-0.01, 0.37) | 0.069 |
| GFAP | 0.18 (-0.04, 0.39) | 0.11 | 0.28 (0.05, 0.48) | 0.017 |
| NFL | 0.26 (0.06, 0.44) | 0.013 | 0.20 (-0.003, 0.39) | 0.053 |
| T-tau | 0.17 (-0.02, 0.36) | 0.083 | 0.16 (-0.05, 0.35) | 0.13 |
| APPa | 0.05 (-0.15, 0.25) | 0.63 | 0.05 (-0.17, 0.26) | 0.68 |
| APPß | 0.03 (-0.17, 0.24) | 0.75 | -0.003 (-0.22, 0.21) | 0.97 |
| YKL40 | 0.25 (0.05, 0.44) | 0.017 | 0.23 (0.01, 0.42) | 0.037 |
| MCP1 | 0.11 (-0.11, 0.32) | 0.32 | 0.12 (-0.12, 0.34) | 0.32 |
| NG | 0.19 (-0.02, 0.38) | 0.072 | 0.16 (-0.06, 0.36) | 0.14 |
| GAP43 | 0.22 (0.03, 0.40) | 0.025 | 0.20 (0.005, 0.39) | 0.045 |

Values are presented as Pearson correlation coefficients and partial Pearson correlation coefficients with 95% confidence intervals (CIs).
Biomarkers were log-transformed prior to analysis to account for skewness.
Missing biomarker data were handled using multiple imputation by chained equations, and results were pooled according to Rubin's rules.
\*Adjusted for iNPH total scale score at baseline (pre-surgery) and age.
**Abbreviations:** Abbreviations: Aβ40, amyloid beta 1–40; Aβ42, amyloid beta 1–42; APPa, amyloid precursor protein alpha; APPß, amyloid precursor protein beta; CI, confidence interval; P-tau, phosphorylated tau; GAP43, growth-associated protein 43; GFAP, glial fibrillary acidic protein; iNPH, idiopathic normal pressure hydrocephalus; MCP1, monocyte chemoattractant protein-1; NFL, neurofilament light chain; NG, Neurogranin; SD, standard deviation; T-Tau, total tau protein; YKL40, chitinase-3-like protein 1.

Regarding the iNPH-scale subdomains, increases in GFAP were significantly correlated with improvement in postoperative gait score and GAP43 with improvement in urinary incontinence. No significant correlations were found between changes in biomarker levels and changes in cognition or balance after surgery. These data are shown in supplementary tables (S4.2-4.5).

### Predictive value of baseline biomarkers

Baseline biomarker levels generally showed limited ability to predict treatment response (Table 5). MCP1 was the only biomarker significantly associated with improvement in the total iNPH scale score, with lower baseline levels associated with greater odds of clinical improvement (adjusted OR 0.45, 95% CI 0.24-0.85; p=0.014). However, predictive performance was modest with an AUC 0.67. The discriminative performance was low to modest for all baseline biomarkers. Consequently, none of the biomarkers demonstrated clinically useful prediction of overall treatment response.

**Table 5.** Association between baseline CSF biomarkers and postoperative improvement (iNPH total scale change score ≥ 5 vs. <5 at follow-up)

| Baseline biomarker | Unadjusted OR (95% CI) | ROC AUC (95% CI) | p-value | Adjusted OR* (95% CI) | p-value |
|---|---|---|---|---|---|
| Aβ 40 | 0.90 (0.60, 1.35) | 0.53 (0.42, 0.64) | 0.60 | 1.06 (0.68, 1.65) | 0.80 |
| Aβ42 | 1.01 (0.69, 1.47) | 0.50 (0.39, 0.61) | 0.97 | 1.09 (0.73, 1.63) | 0.68 |
| Aβ 42/40 ratio | 1.34 (0.63, 2.86) | 0.53 (0.42, 0.64) | 0.45 | 1.09 (0.49, 2.43) | 0.84 |
| P-tau | 0.80 (0.58, 1.11) | 0.59 (0.49, 0.70) | 0.18 | 0.89 (0.62, 1.28) | 0.53 |
| GFAP | 0.77 (0.55, 1.07) | 0.58 (0.48, 0.69) | 0.12 | 0.84 (0.59, 1.19) | 0.33 |
| NFL | 1.08 (0.84, 1.39) | 0.48 (0.38, 0.58) | 0.56 | 1.20 (0.88, 1.63) | 0.26 |
| T-Tau | 0.98 (0.75, 1.27) | 0.52 (0.41, 0.63) | 0.86 | 1.00 (0.75, 1.32) | 0.99 |
| APPa | 0.86 (0.63, 1.18) | 0.55 (0.44, 0.66) | 0.36 | 0.99 (0.70, 1.41) | 0.97 |
| APPß | 0.80 (0.56, 1.13) | 0.56 (0.45, 0.67) | 0.20 | 0.91 (0.62, 1.33) | 0.61 |
| YKL40 | 0.95 (0.59, 1.51) | 0.51 (0.41, 0.62) | 0.82 | 1.08 (0.66, 1.78) | 0.76 |
| MCP1 | 0.44 (0.23, 0.82) | 0.67 (0.57, 0.77) | 0.011 | 0.45 (0.24, 0.85) | 0.014 |
| NG | 0.69 (0.46, 1.04) | 0.59 (0.48, 0.71) | 0.075 | 0.74 (0.48, 1.13) | 0.16 |
| GAP43 | 0.75 (0.49, 1.13) | 0.56 (0.45, 0.68) | 0.17 | 0.83 (0.53, 1.29) | 0.40 |

Statistical analyses were conducted using logistic regression.
Biomarkers were log-transformed prior to analysis to account for skewness.
Missing biomarker data were handled using multiple imputation by chained equations, and results were pooled according to Rubin’s rules.
Results are presented as odds ratios (ORs) of improvement (iNPH total scale change score ≥ 5) vs no improvement per 50% increase in biomarker levels.
\*Adjusted for iNPH total scale at baseline (pre-surgery) and age.
**Abbreviations:** Aβ 40, amyloid beta 1–40; Aβ 42, amyloid beta 1–42; APPa, amyloid precursor protein alpha; APPß, amyloid precursor protein beta; AUC, area under the curve; CI, confidence interval; P-tau, phosphorylated tau; GAP43, growth-associated protein 43; GFAP, glial fibrillary acidic protein; iNPH, idiopathic normal pressure hydrocephalus; MCP1, monocyte chemoattractant protein-1; NFL, neurofilament light chain; NG, neurogranin; OR, odds ratio;
ROC, receiver operating characteristic; T-Tau, total tau protein; YKL40, chitinase-3-like protein 1.

There was a modest association between Aβ40 and Aβ42 and improvement in urinary symptoms. Lower baseline levels of Aβ40 (Adjusted OR 0.63, 95% CI 0.40-0.99; p=0.047), and Aβ42 (Adjusted OR 0.54, 95% CI 0.35-0.83; p=0.005) were associated with less postoperative urinary incontinence. However, the AUC-values indicated low predictive performance for both, at 0.63 and 0.64 respectively. No baseline biomarkers were significantly associated with cognition, gait or balance outcome.

## DISCUSSION

In this prospective longitudinal study of 135 patients with iNPH, we investigated the change in CSF biomarkers following shunt surgery and examined the predictive value of preoperative CSF biomarker levels. The three principal findings of this study are the following: First, shunt surgery was associated with substantial changes in the concentrations of multiple CSF biomarkers. Second, changes in several biomarkers were associated with the postoperative clinical improvement. Third, preoperative biomarker levels showed limited ability to predict treatment response.

Among the investigated biomarkers, the largest postoperative changes were observed for GAP43, NG, P-Tau, and amyloid-related proteins (Aβ40, Aβ42, sAPPα, and sAPPβ). Interestingly, GAP43 and NG share several biological characteristics that make the pronounced postoperative changes observed in the present study particularly noteworthy. Both proteins regulate synaptic function, plasticity, axonal remodeling, and structural circuit adaptation (16, 17).While presynaptic GAP43 drives axonal growth, synapse formation, and injury-induced regenerative sprouting (18, 19), as well as adaptive responses to altered CSF-dynamics in hydrocephalic animal models (20), NG mediates post-synaptic calmodulin signaling within dendritic spines and is essential for synaptic plasticity and long-term potentiation (21) (22) These findings could possibly indicate that successful CSF diversion is accompanied by biological processes related to neuronal remodeling and restoration of synaptic function. Although, it is important to emphasize that the present study cannot establish whether these changes directly contribute to clinical recovery or merely reflect downstream consequences of restored CSF dynamics. Nevertheless, their established roles in neuronal adaptation provide a biological rationale for further investigation in mechanistic studies of disease reversibility in iNPH.

The observed increases in amyloid-related biomarkers postoperatively are also of particular interest. Reduced CSF concentrations of Aβ peptides are well documented in iNPH (23, 24) but their precise role in the pathophysiology and dynamics following CSF diversion is less understood. It has previously been suggested that the low preoperative concentrations of APP-derived proteins in iNPH are the result of reduced production due to hypometabolism particularly in the periventricular areas (25). Possibly the postoperative increase in amyloid-related proteins, seen in this study, reflect increased metabolic activity due to improved perfusion, which has been shown to occur after CSF diversion (26).

The notable postoperative increases in T-tau and P-tau are also of interest. T-tau is generally regarded as a marker of neuronal injury, as elevated CSF concentrations are observed across a range of neurological disorders whereas P-tau is mainly regarded as a key driver in the pathophysiologic process in Alzheimer’s disease (AD) (27). Considering the possible restorative effects of CSF- diversion, the more than two-fold increase in T-tau and P-tau was somewhat surprising since elevation of these proteins are mainly seen in disease states. However, in a previous study of the same cohort where ventricular CSF was sampled via the shunt catheter, we similarly observed substantial postoperative increases in most biomarkers following shunt surgery except, for T-tau (which decreased) and P-tau (which remained unchanged), highlighting an apparent discrepancy between ventricular and lumbar CSF (28). One possible explanation is that restoration of CSF circulation following shunt surgery transiently alters tau kinetics, resulting in increased lumbar CSF concentrations because of slower CSF turnover and protein clearance in the spinal compartment. In contrast, the more rapid turnover of ventricular CSF may lead to earlier normalization of tau concentrations, rendering such transient changes less apparent at the time of postoperative sampling. Although speculative, this hypothesis could be addressed in future studies using serial ventricular and lumbar CSF sampling over an extended postoperative follow-up period.

Overall, the pronounced postoperative biomarker changes observed in this study, together with their association with clinical improvement, might suggest that restoration of CSF dynamics is accompanied by biological processes, at least partially, involved in neuronal recovery and functional reorganization. Whether these biomarker changes reflect mechanisms directly contributing to clinical reversibility or represent downstream consequences of improved CSF circulation (29) cannot be determined from the present study. Nevertheless, the consistent alterations across several markers together indicate that the marked clinical improvement observed after shunt surgery in iNPH is accompanied by measurable changes in biological pathways that ought to be explored further.

We also found that changes in several biomarkers were associated with the magnitude of clinical improvement. Patients with greater increases in GFAP, YKL40, and GAP43 experienced larger improvements on the iNPH scale. These results are partially in line with our earlier study in which the relationship between ventricular CSF-biomarkers and postoperative outcomes were assessed (28). In that study, change in ventricular CSF levels of Aβ40 and Aβ42 and YKL40 correlated with clinical outcome. However, GAP43 did not. Astrocytes are essential for maintaining neuronal homeostasis and white matter function through various mechanisms such as regulation of extracellular ion balance, metabolic support, and cell signaling (30). Consequently, the observed association between postoperative YKL40 dynamics and clinical improvement might indicate that astrocytic responses could be part of the biological processes accompanying successful shunt treatment. The fact that YKL40 changes were associated with clinical improvement suggests that these changes are not solely a consequence of surgery-related inflammation. It is also worthwhile to mention that although the observed correlations between change in CSF-biomarkers and clinical improvement were generally modest, they highlight the fact that longitudinal biomarker measurements can provide more informative insights into disease mechanisms than single baseline measurements.

Although the marked postoperative changes in biomarker concentrations could be due to alterations in cellular homeostasis it is also important to consider whether they instead reflect a mechanical washout effect i.e. increased CSF turnover diluting or clearing existing biomarker pools (31). In this study all biomarker concentrations increased postoperatively, without a distinctive pattern favoring specific markers, which could suggest such a non-specific washout mechanism. However, in a previous study examining both lumbar and ventricular CSF pre- and post shunt surgery, similar findings were noted with marked postoperative increases in amyloid-related proteins but unchanged or decreased levels of inflammatory biomarkers, indicating that the observed changes are not entirely due to increased CSF turnover (25). Similarly, our previous study, on the same cohort, also showed bidirectional biomarker changes in ventricular CSF (28). In that study, levels of amyloid- related proteins, Nfl, YKL40, GAP43 and MCP-1 increased whilst T-tau, GFAP, NG and Aβ42/40 decreased. That study also showed, akin to the present findings, that patients who improved clinically showed more pronounced postoperative changes in several biomarkers compared to non- improved patients. If the biomarker changes were driven entirely by a washout phenomenon, similar magnitude of change would be expected regardless of clinical outcome, which was not the case. Furthermore, the various mechanisms are not mutually exclusive and it is therefore possible that the mechanical effect of increased CSF turnover itself contributes to improved cellular functioning, by reducing the local concentration of potentially neurotoxic species such as amyloid oligomers (32), thereby possibly providing a link between the hydrodynamic and molecular effects of shunt surgery. Nevertheless, the exact mechanisms underlying these findings ought to be further studied as they might aid in the understanding of the pathophysiological processes in iNPH.

Regarding the predictive performance of baseline levels, all biomarkers showed limited value. MCP1, a marker also associated with astrogliosis and cerebral inflammatory processes (33) was the only biomarker significantly associated with overall clinical improvement. Although elevated MCP1 concentrations have previously been reported in iNPH patients (23, 34), it is important to note that its discriminatory performance in this cohort was modest. No biomarker demonstrated sufficient predictive accuracy to support clinical use as a standalone predictor of treatment response. These results align with earlier studies reporting inconsistent or weak associations between baseline CSF biomarkers and shunt outcomes (5, 35–37). One possible explanation is that preoperative biomarker concentrations reflect an underlying biological heterogeneity that does not fully capture the mechanisms determining treatment responsiveness. Another contributing factor may be methodological issues such as selection criteria and varying patient populations across studies, which could affect sampling distributions and influence the likelihood of detecting meaningful between-group differences. Nevertheless, the discrepancy between the pronounced biomarker changes seen in this study and the limited predictive value of baseline levels is noteworthy. While baseline measurements represent a static snapshot, longitudinal changes may more directly reflect the physiological response to restoration of CSF circulation. This distinction may, partly, explain why cross-sectional biomarker studies have yielded inconsistent findings and highlights the value of prospective longitudinal designs.

The strengths of this study include its prospective design, the relatively large cohort, the standardized clinical assessments, and the availability of paired pre- and postoperative CSF samples. Several limitations should also be acknowledged. The observational design limits causal inference regarding the biological mechanisms underlying biomarker changes. Importantly, although the observed biomarker changes were pronounced, the present study cannot fully elucidate the exact mechanisms and how they related to the actual clinical response, i.e whether they reflect restoration of a disease state that leads to improvement or merely a coexisting non-causal phenomenon. Also, the study lacked a control group, and multiple biomarkers were examined, increasing the risk of type I error. Furthermore, although statistically significant associations were observed, several effect sizes were modest and should be interpreted cautiously.

In conclusion, shunt surgery was associated with substantial longitudinal changes in multiple CSF biomarkers, several of which correlated with the degree of clinical improvement. These findings suggest that biomarker dynamics may reflect biological processes related to treatment response and reversibility in iNPH. In contrast, baseline biomarker levels demonstrated limited ability to predict clinical outcome. Longitudinal biomarker studies may therefore provide a valuable framework for improving our understanding of disease mechanisms and treatment-related biological effects in iNPH.

## Data Availability

The data produced in the present study are not publicly available due to ethical and legal restrictions

